# The effect of free school fruit on academic performance: a nationwide quasi-experiment

**DOI:** 10.1101/2022.12.08.22283247

**Authors:** Torleif Halkjelsvik, Elling Bere

**Affiliations:** Norwegian Institute of Public Health

**Keywords:** fruit, school, academic performance, quasi-experiment

## Abstract

In past research, higher intake of fruit has been associated with better academic achievement. Can the provision of one piece of fruit every school day improve children’s academic performance? In Norway, the government required all lower secondary schools to provide fruit to their pupils from 2007 to 2014. The policy also covered schools with combined elementary and lower secondary education (1st to 10^th^ grade), but not ordinary elementary schools (1^st^ to 7^th^ grade). This differentiation, in combination with administrative data on test scores before, during, and after the law was enforced, created a nationwide quasi-experiment. Population register data on parents’ sociodemographic characteristics allowed for targeted analyses of a subsample with lower grades and lower fruit intake (boys of low socioeconomic status). In pre-registered analyses, we found no evidence that exposure to the free school fruit policy improved academic performance in the subsample or the entire population of Norwegian pupils. The free fruit policy coincided with a slight decline in performance among pupils covered by the policy. In a Western country with low levels of food insecurity, a policy that required schools to provide free fruit to pupils did not improve learning and may even have interfered with learning.

**Significance Statement:** The intake of fruit is believed to be beneficial for children’s concentration, and research has linked a higher intake of fruit to better academic performance. During a national policy that made it mandatory for some types of elementary schools to provide one daily piece of fruit to every pupil, we did not observe any beneficial effects on learning. On the contrary, the policy coincided with a slight performance decline. Even for low-socioeconomic boys, who have lower-than-average grades and a lower intake of fruit, there was no improvement in academic results associated with the policy. We speculate that the policy may have required resources that otherwise would be used for teaching or teaching-related administration.

## Introduction

The school provides a convenient arena for interventions and measures targeting childhood nutrition. In developed countries, free school meals (1) and improved school meal quality (2) have been reported to improve academic performance. Long-term beneficial outcomes such as higher educational attainment and higher adult income have been reported as potential effects of the current Swedish school meal program (3), the historical Norwegian Oslo breakfast (4), and the US National School Lunch Program (5). The literature on healthy eating at school and academic performance has mainly focused on broad nutritional interventions relating to meals such as lunch and breakfast. Few, if any, studies have systematically assessed the impact of providing more specific types of food to school children. The present study exploits the differential impact of a nationwide policy to assess whether providing fruit to school children can affect their academic performance.

The government’s argument for providing free fruit to school children was that a healthy diet would contribute to better learning (6, 7). Apart from lay beliefs and expectations by politicians and authorities regarding the benefits of fruit, there are several more scientific links between the consumption of fruit and the potential for improved learning.

In general, fruit is considered an important component of recommended healthy diets (8), and it may contribute to preventing a range of chronic diseases (9) that can hamper social and cognitive functioning. Youth consume many ultra-processed foods that might be negatively linked to cognition and learning (10), and fruit may replace such unhealthy food (11).

Fruit contains several basic nutrients, as well as other compounds with potential benefits beyond basic nutrition. Particular nutrients and secondary metabolites found in fruits act on molecular systems and cellular processes that are vital for maintaining cognitive function (12, 13), also for young people (14). Gut microbiota is considered important for cognition (15), and eating fruit contributes to a healthy gut microbiota (16). In research on fruits, polyphenols (17, 18) have gained great attention. Flavonoids (a polyphenol, abundant in fruits) might benefit cognitive outcomes within an acute time frame of 0–6 hours (19). Fruit contains sugar, which in some (but not all) studies has been found to have acute beneficial effects on cognition (20).

Fruit intake has been associated with better cognitive performance (21). Fruit might also increase concentration in school children (22), which may reduce negative behaviors (off-task, out-of-seat) that impair the learning environment for the other children in the classroom.

In several international studies, fruit intake and diet quality in general have been associated with better academic performance (23-25). This has also been shown in Norway. A recent study of 15–17-year-olds reported that among girls, 40% of those with high academic achievement ate fruit daily, while the figure was only 25% for those with low academic achievement (26).

As studies of fruit and academic performance are typically observational and cross-sectional, we do not know if fruit consumption among children causes better academic performance. There is a social gradient in health and health behaviors, and also in fruit intake in adolescents in Norway (27). Healthy eating is also correlated with more physical activity and healthy sleep habits, behaviors that themselves might affect academic performance (28). Furthermore, intelligence, which is one of the main determinants of academic performance, might also be related to dietary choice and fruit intake (29).

Such problems of confounding variables represent a key problem in nutrition epidemiology, and fewer but larger (mega) trials have been suggested as a solution to achieve better answers to the most important questions (30). However, such trials are difficult to conduct (31), and therefore rare. Due to the nature of the implementation of a free school fruit policy in Norway (32), the current study circumvents some of the methodological problems of past research on fruit and academic performance.

In 2007, the Norwegian Government required that all children and adolescents in schools with lower secondary education (grades 8 to 10, age 13 to 16) were to receive (free of charge) a piece of fruit every school day. In 2008 this was required by law after a proposal from the Ministry of Education (6). The law was repealed in 2014 because of high costs and consideration of the autonomy of schools and municipalities (33). This means that the policy was in effect from the school year that started in autumn 2007 (the school year 2007/2008) to the end of the school year that started autumn 2013 (the school year 2013/2014). The policy included all schools with lower secondary education, which also meant that elementary pupils in combined elementary and lower secondary schools (20% of elementary pupils in Norway) were covered. Children in ordinary elementary schools (80%) were not eligible for free fruit and is used as a control group. There is a tradition for bringing packed lunch in Norway, and school-provided breakfast or lunch is not common in elementary school. This means that for the majority of the pupils eligible for free fruit, the piece of fruit was the only food item provided by the school.

The consumption of fruits in Norway has traditionally been low, and before the free school fruit implementation, it was argued that Norwegians ate less than half of what is recommended by the government (34-36). According to earlier reports, the free fruit policy increased average fruit intake by approximately half a portion (about 30%) among the children attending schools covered by the policy (37, 38), and it increased the proportion eating fruits daily by 10 percentage points (from 57% to 67%) (39). Furthermore, it has been reported that it reduced unhealthy snack consumption in children from families with low socioeconomic status (11).

To estimate the potential effect of the policy on learning outcomes, we categorized schools into treatment and control schools based on their status as combined (elementary and lower secondary education) or divided (elementary education only). We obtained records of the pupils’ academic performance on objectively graded national tests in Mathematics, Reading, and English. The tests are carried out approximately a month into the school year in the 5^th^ (elementary school), 8^th^ (lower secondary school), and 9^th^ grade (lower secondary school). In addition, we also received school-level exam results for 10^th^ grade.

The policy lasted 7 years, which gives a graded exposure of free fruit in elementary school from one to four years before the 5^th^ grade test, and one to seven years before tests in the lower secondary school. The treatment (eligibility for free fruit) refers to the provision of free fruit during *elementary school*, because during lower secondary education (8^th^ to 10^th^ grade) either all pupils (school years 2007/08-2013/14) or none (school years before 2007/08 and after 2014/15) of the pupils received free fruit the previous year.

We were particularly interested in pupils with potentially low intake of fruit and lower-than-average performance. Boys are found to eat less fruit than girls, and in comparison with pupils of higher socioeconomic status, pupils of lower socioeconomic status eat less fruit (40). Some groups of immigrant origin have low consumption of fruit, whereas the traditional food of other groups includes a high amount of fruit and vegetables (41). Thus, non-immigrant boys of lower socioeconomic status represent a relatively homogenous demographic group of particular interest. In addition to the targeted analyses of this subsample, we assessed the potential impact of the policy on all registered pupils.

## Results

### Descriptive Results

Figure 1 presents all registered scores of the three 5^th^ grade tests for boys with low socioeconomic status and the full sample of all 5^th^ graders in Norway. The number of years the pupils had received free fruit in the treatment schools is indicated in the upper horizontal axes. Exposure to the free fruit policy in control schools was 0 for all years. See Supplemental Materials (Figures S1 and S2) for standardized scores. Although the control and treatment schools develop in parallel, there appears to be a larger discrepancy during the years of receiving free fruit.

**Figure 1.**
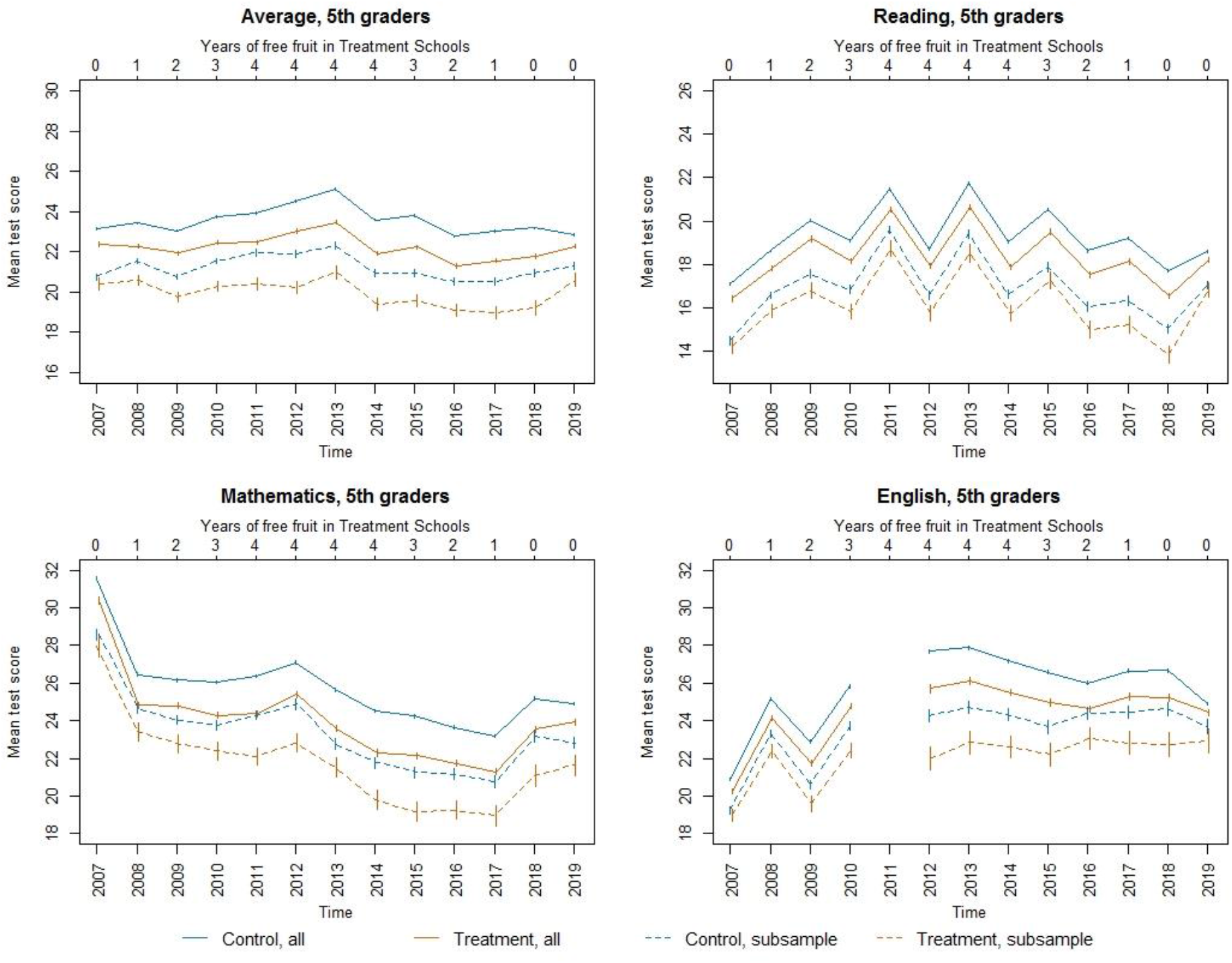
Averages of original scores for all 5^th^ graders and subsample of non-immigrant boys with low socioeconomic status. Error bars are 95% confidence intervals of means (N per year by test from 685,373 to 741,975 in the full sample and 64,261 to 70,663 in the subsample).

### Main Analyses

Regression analyses estimated differences in test scores as a function of exposure to the free fruit policy, controlled for school-specific differences and differences between years. The exposure to the fruit policy was estimated by separate Phase-in and Phase-out terms. There was not a gradual roll-out of the policy, but a gradual accumulation and decrease in the number of years the pupils were potentially exposed to the policy. A third term controlled for having fruit in the test year (tests are carried out at the beginning of the semester). A priori, we chose to focus on the Phase-in term (See pre-registration https://osf.io/uefjp).

None of the pre-registered analyses on 5^th^ graders suggested any positive impact of the policy on test scores (Supplementary Materials, Table S1). Contrary to our expectations, a one-year increase in the exposure to the free fruit policy was associated with a decrease in test scores of 0.18 (0.02 standardized points) in the subsample and 0.14 (0.01 standardized points) in the full sample (See Table 1). Exposure to any number of years of free fruit was associated with a decrease in test scores of 0.66 points (0.08 standardized points) for the subsample and 0.50 (0.06 standardized points) for the full sample. The coefficients for the Phase-out, which measured results on tests carried out when the policy was no longer active (exposure to free fruit but not the most recent year), were also negative, but of lower magnitude and high statistical uncertainty.

**Table 1.**
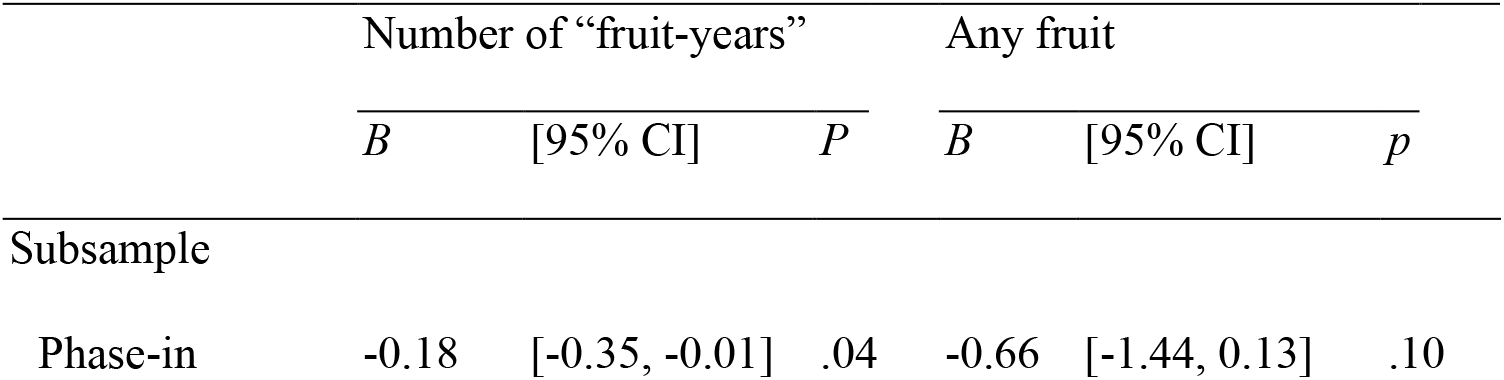
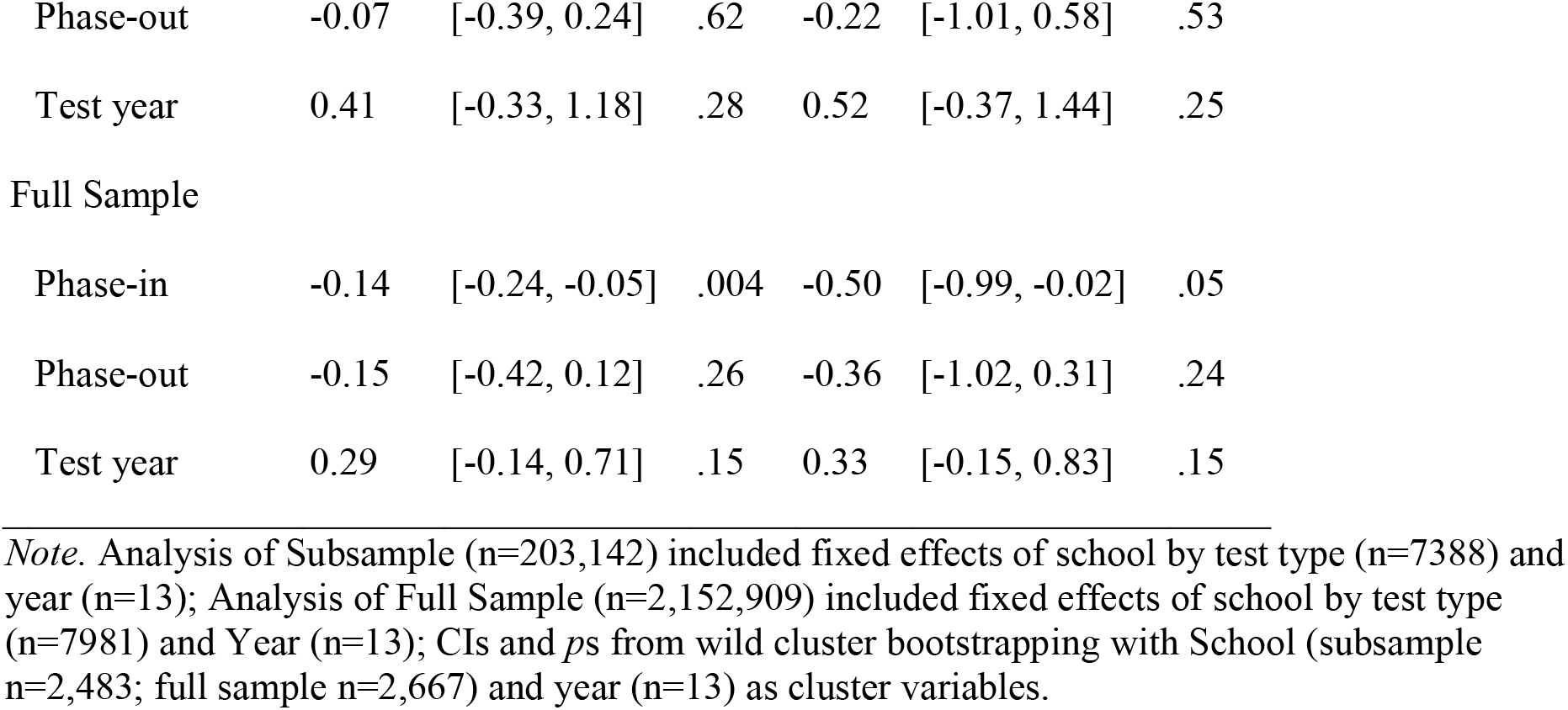
Regression coefficients (B), confidence intervals (CI), and p-values (p) for analyses of the Subsample and Full Sample, 5^th^ grade tests.

In covariate-adjusted analyses, we included population registry data on the pupil’s sex and both parents’ birth country, income, and education. In addition to this pre-specified covariate-adjusted analysis, we ran models that additionally controlled for the proportion of pupils exempted from the tests (or otherwise missing), school’s number of pupils registered for the test, and municipality centrality/rurality by year. These analyses gave a similar pattern of results as the main analyses and are presented in the Supplemental Materials (Tables S2, S3, and S4).

In addition to the 5^th^ grade tests, we pre-specified analyses on 8^th^ grade tests. Unfortunately, data that identified the pupils’ elementary schools (school the previous year) were not available in the registry for 8^th^ grade tests in 2007 and partly missing subsequent years (see Supplementary Materials, Table S5). An analysis on the available data did not detect any positive effect of the policy and showed tendencies of a negative association between the number of years of free fruit and test scores, B = -0.17 [-0.32, -.0.03], *p* = .02 for the subsample and B = -.0.07 [-0.14, 0.01], *p* = .08 for the full sample (Supplementary Materials, Table S6).

### Exploratory Analyses

To aid the interpretation of the unexpected pattern of results, we acquired additional data on 10^th^ grade exams. The average exam scores for 10^th^ graders in the Treated and Control schools are presented in Figure 2. Note that school type in 10^th^ grade is used as a proxy for school type in elementary school (combined school in 10^th^ grade = combined elementary school [treatment group], divided lower secondary school = divided elementary school [control group]). The pre-policy developments of the treatment and control schools were fairly similar, with parallel trends, but they appeared to slightly diverge when cohorts were differentially exposed to the free fruit policy. A regression analysis similar to the main analysis suggested negative longer-term effects corresponding to a 0.1 decrease in school points (or 0.01 grade points; school points are grades multiplied by 10) for each year of exposure while the policy was in place (Phase-In = -0.13, 95%CI[-0.24, -0.02], *p* =.03) and for each year of exposure in the past, after the policy was repealed (Phase-Out = -0.10, 95%CI[-0.15, -0.04], *p* = .002; See Supplementary Materials, Tables S8 and S9).

**Figure 2.**
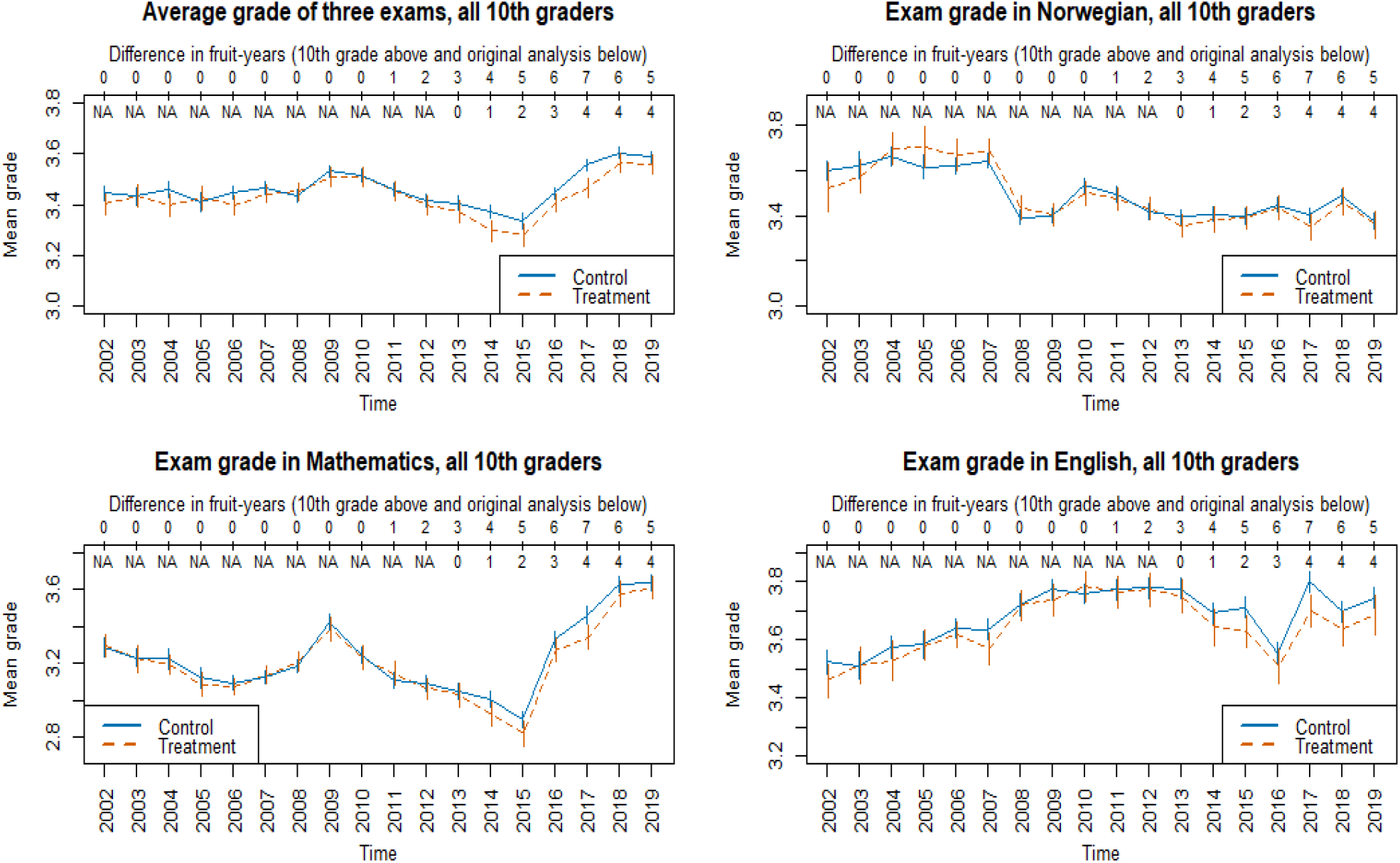
The development in 10^th^ grade exam scores before and during the study period. Error bars are 95% confidence intervals of means. Grade range from 1 (worst) to 6 (best).

Using the pupils’ lower secondary school as a proxy for elementary school also allowed us to use more of the 8^th^ grade data. An alternative analysis of the 8^th^ grade data using school in 8^th^ grade as a proxy for elementary school did not detect any positive or negative effects, *p*s > .3 (Supplementary Materials, Table S7). Similar analyses of 9^th^ grade national tests (only available years 2010-2019) revealed negative estimated effects of exposure to the policy for both the phase-in and phase-out terms (see Supplemental Materials, Table S10). Supplementary analyses using aggregate data (to avoid the potential problem of anti-conservative inferential statistics caused by few time clusters and large cross-sectional samples, see e.g. (42)) showed relatively consistent effect size estimates corresponding to a decrease of approximately 0.1 original points (0.01 standardized points) for each year of free fruit exposure (Supplementary Materials, Table S11).

The negative estimated effect is likely not due to simple time by treatment status confounding, because the Phase-out term (fruit exposure but not the previous year) of the 10^th^ grade data starts at 2015, whereas the Phase-in term (fruit exposure while policy is active) of the 5^th^ grade data ends at 2014, and both these indicate a negative effect of increased exposure to the free fruit policy. Cohort by treatment status confounding is more plausible as indicated in Figure 3, where the relative scores of treatment schools are sorted by the normal birth year of pupils in the cohorts.

**Figure 3.**
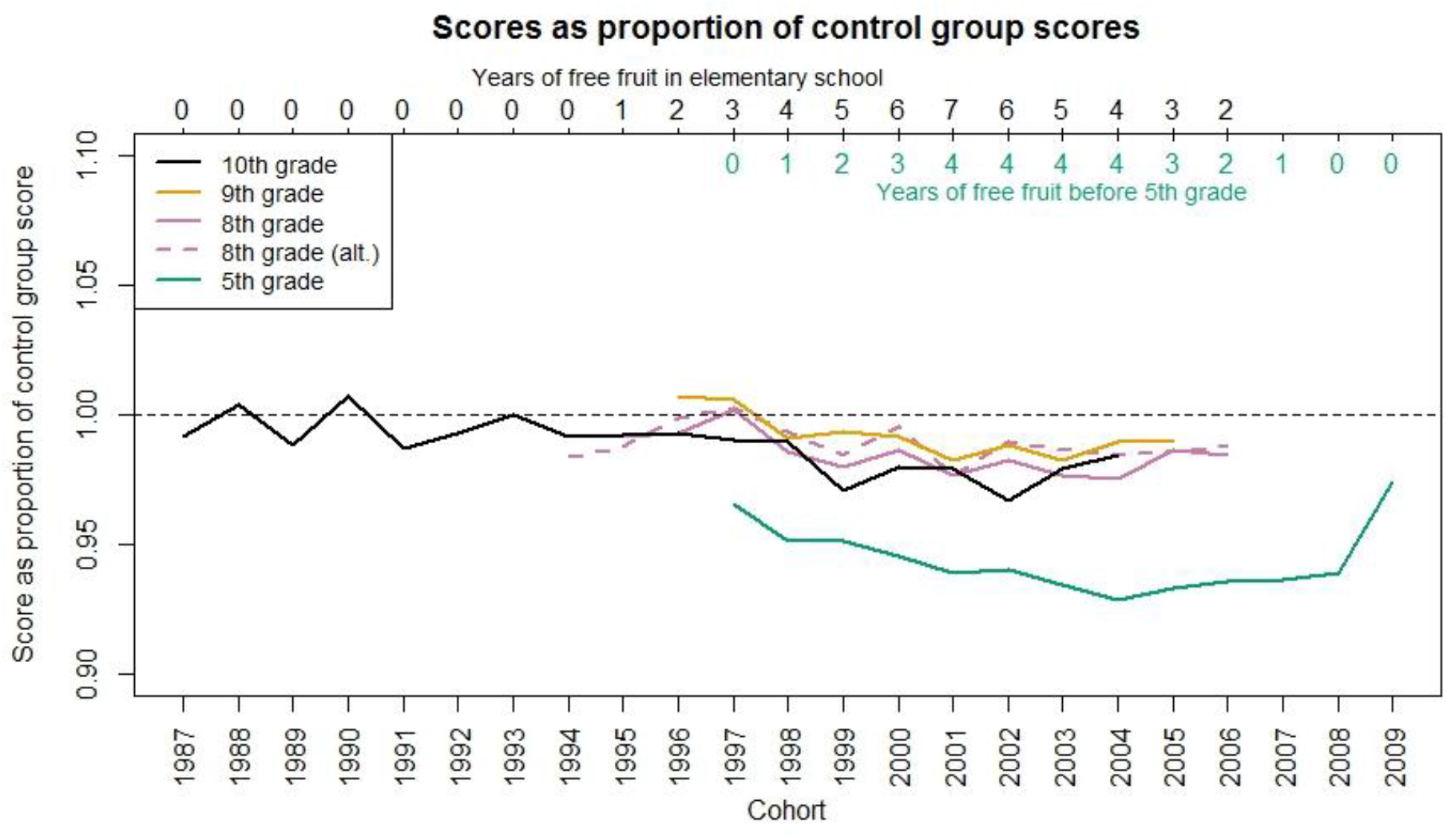
Relative scores of treatment schools sorted by cohort (across tests taken at different times). Abbreviations: alt.=alternative categorization of treatment status for 8^th^ grade based on current instead of previous school (this also applies to 9^th^ and 10^th^ grade data).

To account for potential factors that differentially affect cohorts in treatment and control schools, we analysed within-person changes from 5^th^ to 8^th^ grade in parts of the data. In the cohorts born 2001 to 2004, pupils in the treatment group all received 4 years of free fruit before the 5^th^ grade test, but a decreasing number from 7 to 4 years of free fruit before the 8^th^ and 9^th^ grade tests. These analyses were consistent with the original analyses (Supplementary Materials, Table S12). In analyses that used all available within-person data (cohorts 1997 to 2006) and a single term for all variation in fruit exposure (assuming an identical effect of increased and decreased exposure before 5^th^ grade and before 8^th^/9^th^ grade), the estimates were close to zero (Supplementary Materials, Table S13).

## Discussion

In Norway, during the period 2007 to 2014, schools with combined elementary and lower secondary education were required to provide one piece of fruit to all pupils every school day. Using ordinary (divided) schools as a control group, we modeled the impact of the policy on test scores. We did not find evidence for a beneficial effect of the policy on academic performance. In pre-registered analyses, the estimated effects tended to be negative both in a sample of all registered Norwegian 5^th^ graders and in a subgroup of pupils known to have a low intake of fruit. However, in the latter subgroup and in analyses of 8^th^ graders, the statistical support for a negative effect was weak. Explorative analyses that used a proxy for treatment status (current school type instead of elementary school type) detected no association between performance and level of exposure to the free fruit policy in 8^th^ grade, but negative associations in 9^th^ and 10^th^ grade. Within-subject analyses on parts of the data gave estimates that were closer to zero.

The free fruit policy was implemented to improve learning outcomes by improving diets. Large proportions of the personnel responsible for the administration of fruit locally at the schools (e.g., teacher, principal, janitor, or other staff) indicated that the program was beneficial for pupils’ concentration (49%) and the social environment (66%); few indicated that it was not beneficial (5% and 6%, respectively) (43). Municipalities (i.e. the school owners) informally reported better concentration and learning outcomes among their pupils during the initial year of the policy, and in a public hearing, several institutions pointed out that such direct beneficial effects are supported by past research and experience (6). However, the literature is scarce on school fruit programs and academic performance.

Based on the broader literature on school meals, it is not unreasonable to expect a positive effect of food programs on learning. A literature review of studies from developed countries indicates that universal free school meals that included lunch improve academic performance (1), but results from studies that only studied universal free school breakfast were mixed. Positive outcomes of school breakfast have been reported for concrete behavior observed in the classroom, such as less off-task and out-of-seat behavior (44), and on cognitive performance, e.g. on memory and attention tasks (45). A recent US study indicates that academic performance increases when meals are provided by vendors that focus on healthy food, such as whole grain, vegetables, and fruit (2). Thus, from a general and very broad perspective, one can argue that food provided by the school has the potential to improve learning outcomes.

Why then did the school fruit policy not improve learning? The main links in a potential causal chain between the provision of free fruit at school and higher scores on the national tests can be considered as follows: A free fruit policy increases pupils’ exposure to fruits at school. Pupils’ fruit intake at school will then increase, and the pupils’ diet will improve. Improved nutritional status will improve learning, which is assumed to be reflected in higher test scores.

We do not know the exact individual exposure of the free fruit program, but as presented in the introduction, independent studies suggest that the fruit policy had a positive impact on the pupils’ consumption of fruit. This gives us two questions for discussion: Was the impact on diet quality too small? Is the previously shown association between fruit intake and academic achievement in the literature noncausal?

The effect of school meals on cognitive and academic performance has historically been easier to establish in children whose nutritional status is compromised (43, 44). Food insecurity among pupils of low socioeconomic status is associated with a range of behavioral and cognitive problems (46, 47). It could be that school-based nutritional interventions and policies mainly benefit the academic performance of demographic subgroups that experience food insecurity. Some recent US studies on universal free school meals suggest that school meals can improve academic performance across socioeconomic groups (48-50), but note that the pupils in the US free school meals programs may represent a selective sample, as the program is only available for schools with a high number of pupils defined as poor.

As food insecurity in Norway is rare, but still present (51), the results from past research may not be generalizable to the current Norwegian context. The diet in Norway, including the children’s diet (40), is considered to be reasonably good according to the last dietary surveys—although there is room for improvement (52). The quality of Norwegian diets varies according to socioeconomic status. Men and individuals of low socioeconomic status eat less fruit than women and individuals of high socioeconomic status (53). Thus, if the provision of fruit is to increase the nutritional status of pupils, this should be more likely in a subgroup of boys with low socioeconomic status (who also have relatively low scores on academic tests and thus greater potential for improvement). Our targeted analyses on this demographic group did not reveal any beneficial effects on test scores, and the estimated negative effects appeared to be of similar magnitude as the effects across all pupils.

Several studies have documented associations between diet and academic performance, also specifically between fruit intake and performance (23-25). The results of the current study do not reflect this, in spite of other studies suggesting that the fruit policy increased the proportion of lower secondary pupils eating fruit daily from 57 to 67% (39). Thus, it appears unlikely that the cross-sectional association reflects a causal effect. This is also suggested by a recent study that approaches the question of causation using Mendelian randomization (54). The study reported that different dietary patterns influence performance in various school subjects, but fruit (or vegetables) did not represent a significant component in any of the main patterns found to be beneficial.

In sum, one reason for not finding an effect of the policy is that the association between fruit intake and academic achievement is not causal. Another is that the treatment is too weak, because the provision of free fruit only has a minor impact on the overall diets of the pupils, and/or because the baseline dietary status of Norwegian pupils is already reasonably good.

If we assume that the negative tendency on test scores in the present study reflects a causal effect of the policy, it is theoretically difficult to argue that an increased fruit intake decreased academic performance. A more plausible, although speculative, explanation might be that the organization of the program had negative side effects in terms of reduced time for teaching and/or other necessary administration.

The Norwegian school fruit program was criticized in the media for being hasty and disorganized in the beginning (55). The proposition to repeal the free fruit policy pointed to concerns about how the distribution of fruit was organized (e.g. challenges relating to personnel and logistics) (33). Most of the persons responsible for the general administration of the fruit at the schools (teacher, principal, janitor, or other staff) reported spending between 10 and 60 minutes each week (56). If additionally, the teachers were involved in distributing the fruit to each pupil in class, or if the fruit was provided as a snack that required an extra break, a small disruption or reduction in teaching may be possible. A recent Norwegian qualitative study reported that a newly extended national school-milk subscription scheme added to the teachers’ time burden (57). Reduced teaching time was also reported in a study on the implementation of a mid-morning breakfast program in Peru (58). As the direction of our results were unanticipated, we do not wish to emphasize this post hoc explanation.

A strength of the study is that the decision to provide fruit to combined schools was related to the administrative status of the schools, not to other characteristics of the schools. However, the school types still differ in several respects (see Methods, Table 2). Pupils in the treatment schools have parents with less education and lower income than pupils in the control schools. The treatment schools are located in more rural municipalities, and for 5^th^ grade tests, they scored lower on the tests also in the year before the policy was implemented (see Figure 2). If these differences reflect stable characteristics of the schools (i.e. the typical demography of the pupils), they are by design accounted for in the analyses. If the composition within schools changes across years or cohorts, this is at least partly accounted for in the covariate-adjusted analyses. However, there may be unmeasured compositional differences that drive the effects. The exploratory within-subject analyses can in principle account for such confounding, yet, we do not consider these analyses as the most trustworthy. Only parts of the data could be used, there were few clusters to account for potential heterogeneity in change scores, and the analyses assume equal influence of fruit in different phases and at different ages. Similar to the main analyses, the within-subject analyses detected no beneficial effect of the policy on learning and tended more towards negative than positive estimates.

**Table 2.**
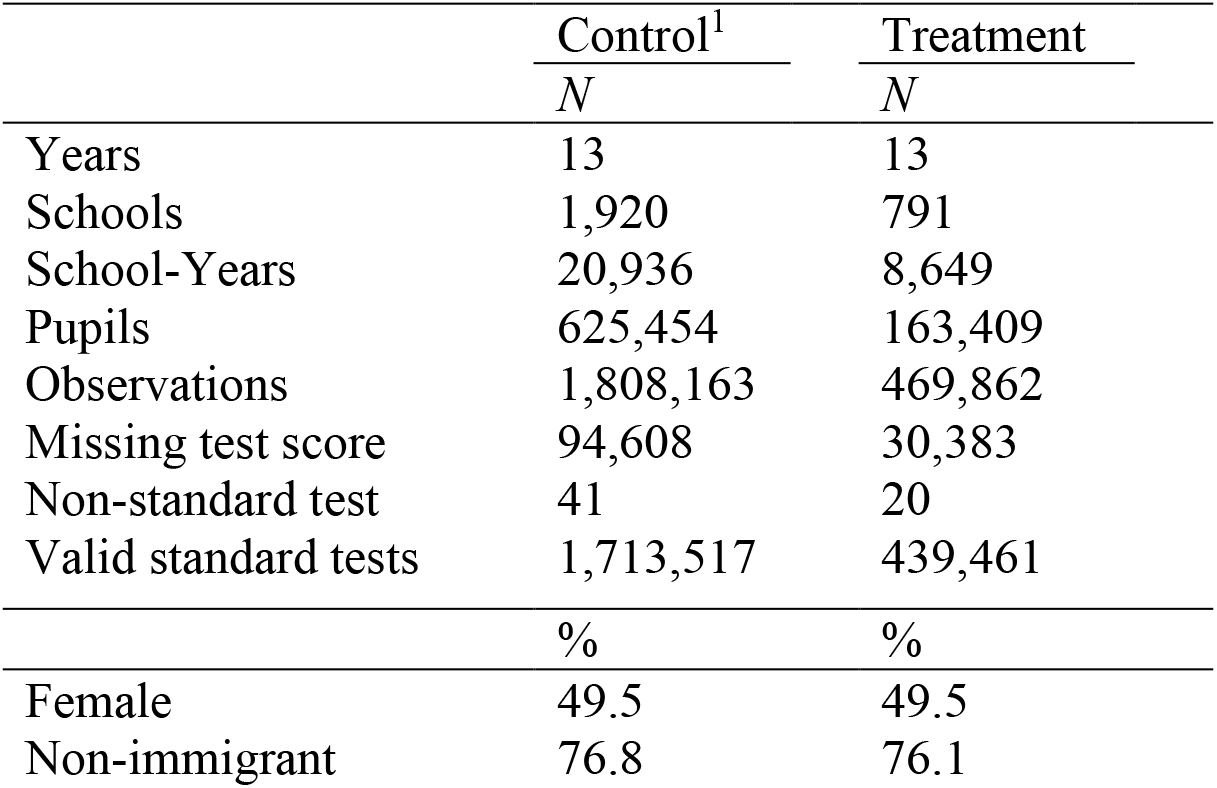
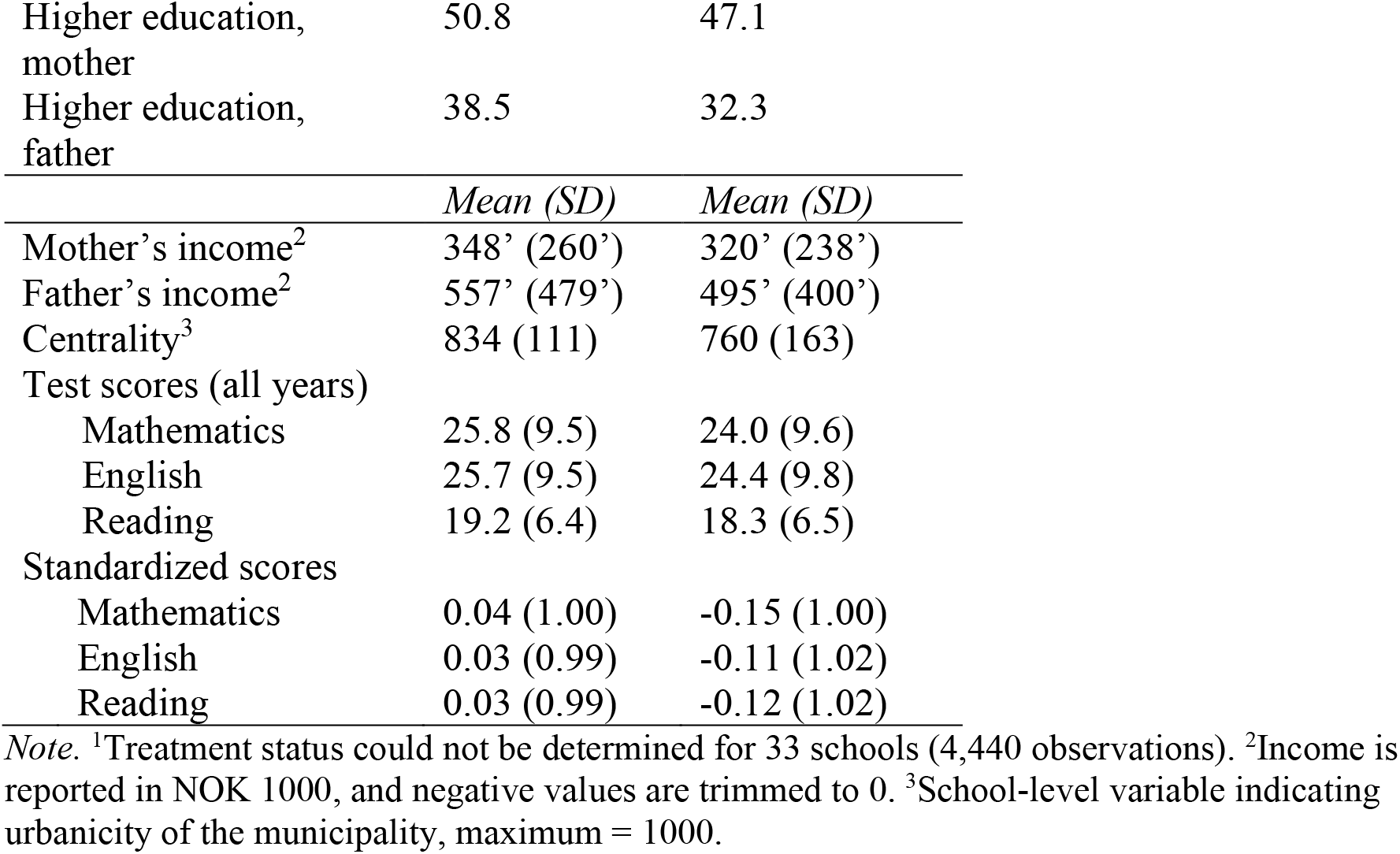
Descriptive statistics for observations in control and treatment schools, 5^th^ grade tests.

The present analyses are like intention-to-treat analyses, where we use data from all pupils regardless of consumption. In other words, we estimate the effects of a policy, not directly the effects of eating fruit. Relatedly, we do not have school details regarding other relevant programs running before, during, or after the free fruit policy. Few elementary schools have school meal programs in Norway, but the number is increasing. In 2013 approximately half of the combined schools, but only one-tenth of the ordinary elementary schools offered paid services such as school canteen, most of the schools offer subscription-based (paid by the parents) milk and other beverages, and 57% of the ordinary elementary school leaders reported to have a kind of fruit arrangement (mostly a subscription program) (59). However, due to low participation, fruit subscription has a limited impact compared to a free fruit program (60). For example, in 2006, just before the implementation of the free fruit policy, 41% of eligible schools participated in a national subscription program, and 28% of pupils at participating schools subscribed, reaching only 12% of potential pupils (61). As the subscription-based fruit programs were available to all schools before the free fruit policy, to the ordinary elementary schools during the time of the policy, and again to all schools after the abolishment, this is in principle not a problem for identifying the effect of the free fruit policy. Furthermore, the pupils that subscribed were typically of high socioeconomic status and consumed more fruit and vegetables in the first place compared with non-subscribers (60).

## Conclusion

In Norway, a Western country with low food insecurity but lower-than-recommended fruit and vegetable intake, the government required some school types to give pupils one piece of free fruit every school day. Although informal evaluations (e.g., satisfaction and perceptions of pupils’ concentration) were generally positive, register data reveal no or even a negative effect of the policy on learning.

## Methods

### Participants and Data

The full dataset on national tests (Mathematics, English, and Reading) consisted of all registered pupils in Norway in 5^th^ and 8^th^ grade in the years 2007 to 2019 (790,242 pupils in 5^th^ grade and 798,869 in 8^th^ grade) and all pupils in 9^th^ grade in the years 2010 to 2019 (only Mathematics and Reading). Exam data in 10^th^ grade is at the school level (N=954; approximately 90% of Norwegian pupils) from the years 2002 to 2019 (Mathematics, English, and Norwegian). The pupils were categorized into treatment and control according to the status of the pupil’s elementary school, which was the school registered at the test in 5^th^ grade, and the registered previous school at the 8^th^ grade test (8^th^ grade is the first year in lower secondary school). For the 9^th^ and 10^th^ grade data, we did not have access to the pupil’s elementary school, and we therefore used their lower secondary school status as a proxy for elementary school status. In 8^th^ grade, 88% of the students were recorded with the same school status (combined versus divided) for previous and current school. Descriptive statistics according to treatment status are reported in Table 2 for the 5^th^ grade test and the Supplemental Materials, Table S5, for the 8^th^ grade test.

The criteria for inclusion in the targeted analysis of pupils known to have a low intake of fruit were: a) male, b) non-immigrant status, c) median household income below the yearly median, and d) none of the parents registered with completed higher education.

The study was approved by the Data Protection Officer at the Norwegian Institute of Public Health. Informed consent was not required due to §8 in the Personal Data Act.

### Deviation from the Pre-Registration

Analyses were pre-registered: https://osf.io/uefjp. Due to delays in the project, we received one extra year of data (i.e. 2019). Original pre-registered analyses without the extra year are included in the Supplemental Materials (Table S14). We planned to rely on the inferential statistics from Linear Mixed Models because these models provided more conservative results than the cluster-robust Fixed Effects models in simulations (see pre-registration). However, due to non-convergence and modeling issues (heterogeneity and difficulties in analyzing standardized scores due to singular fit and non-convergence), we focus on the cluster-robust Fixed Effects models. The inferential statistics of the Linear Mixed Models were not more conservative (Supplemental Materials, Text S1 and Tables S15 and S16).

We specified three different impact models (See Supplemental Materials, Table S17). The first impact model was our a priori best guess of a diminishing impact according to number of years of free fruit received (one year coded 1, two years coded 1.5, etc.), based on the idea that free fruit could establish better habits during initial years or that a potential beneficial effect would be due to the elimination of some nutritional deficit. In the absence of such positive effects, the diminishing impact model is best considered as an arbitrary scale of exposure, and its coefficients are not directly interpretable. Therefore, we included this model only in the supplementary materials (the results were consistent with our second impact model). We focused on our second impact model, which was pre-specified as a model to interpret the magnitude of potential effects. This impact model was based on the actual number of years of free fruit (one year = 1, two years = 2, etc.). We additionally report results from a third pre-registered impact model that used any fruit versus no fruit (one year = 1, two years = 1, etc.), but this model could not be used for the original 8^th^ grade analysis due to missing data the first year (when exposure was 0). Pre-specified Bayes Factors are reported in the Supplemental Materials (Text S2).

### Statistical Analyses

The Fixed Effects regressions were estimated by OLS with the user-written function ‘reghdfe’ (62) in Stata 15.0. Potential heteroscedasticity and dependence within time and school clusters were accounted for by two-way cluster-robust inference. The inferential statistics were either based on the default cluster-robust standard errors or wild cluster bootstrapping (999 replications; null imposed) with the user-written function ‘boottest’ (63), as specified in the tables. The analyses included fixed effects of year and school by test type (Mathematics, English, Reading).

To illustrate the variation in test scores over time, and for communicative purposes, we chose to focus on the original scores of the tests, but we report the main results of standardized variables (M = 0 and SD = 1 within each test type by year) in text and further results on standardized scores are referred to in text and included in the Supplemental Materials.

## Data Availability

The data is not publicly available but may be obtained by application to Statistics Norway (only for researchers affiliated with approved research institutions). See https://www.ssb.no/en/omssb/tjenester-og-verktoy/data-til-forskning

## Acknowledgments

We would like to thank Jonas Kinge, Kjetil Telle, and Anders Skrondal for valuable inputs regarding the analyses, and Arnfinn Helleve and Knut-Inge Klepp for valuable comments on the manuscript.

