## Supplementary material for "The effect of free school fruit on academic performance: a nationwide quasi-experiment"

### Supplementary Information

#### Supplementary Text S1: Linear Mixed Models

Results for Linear Mixed Models are reported in Table S15 (see Table S16 for model example). In addition to the (potential) exposure variables (Table S17), the treatment group indicator, and their interactions, the model controlled for test type, treatment by test type, and included the following random effects: participant intercepts, school intercepts, time intercepts plus random slopes for treatment status (i.e., heterogeneity across time that differs between control and treatment schools at the group level), and school by time intercepts.

We used the default settings in the R (1) packages ‘lme4’ (2) and ‘lmerTest’ (3). The majority of the models failed the default convergence criteria (based on absolute gradients). However, except for one model, the relative gradients were acceptable (<0.0002) and these models converged when using an alternative implementation of the optimizer. Consequently, we only treated one model as truly non-converged (see Table S16). The models were slightly misspecified in terms of the variance structure (there is lower variance for Reading than for the other tests, see Table 2), but we considered the models to be acceptable approximations. We did not succeed in running more complex models in other available software (e.g., due to memory limitation and software crash).

**Supplementary Text S2: Bayes Factors**

As an alternative way of quantifying statistical uncertainty, we calculated Bayes Factors (BFs) for the main analyses (see (4)). These are included in the Supplementary Materials (Table S18). They were pre-specified as a comparison between the likelihood of a null effect (exact null) and the likelihood of a distribution of positive effect sizes. The distribution was a half t-distribution with 3 degrees of freedom. This distribution is close to a half-normal distribution (folded normal distribution) but with a slightly longer tail to accommodate potentially larger effect sizes. The mode of the distribution was 0 and the standard deviation was 0.03 for the continuous impact models (linear and non-linear effect) and 0.05 for the dummy impact model. These ranges of effect sizes cover the typical small effects observed in large RCTs on school interventions (see (5)). The BFs supported the conclusion that a positive impact of the policy is unlikely given the data and the models. We did not report these one-sided BFs in the main text because they are easily misunderstood in the context of negative results (they compare the likelihood of the null with the likelihood of a distribution of positive effect sizes, not negative).

*Table S1. Regression coefficients (B), confidence intervals (CI) and p-values (p) for analyses of raw scores and standardized scores, Selective and Full Samples, 5th grade tests.*

| | Diminishing Impact | | | Number of fruit-years | | | Any fruit | | |
|---|---|---|---|---|---|---|---|---|---|
| | *B* | [95% CI] | *P* | *B* | [95% CI] | *p* | *B* | [95% CI] | *p* |
| Selective Sample | | | | | | | | | |
| Original Scale | | | | | | | | | |
| Phase-in | -0.39 | [-0.81, 0.00] | .05 | -0.18 | [-0.35, -0.01] | .04 | -0.66 | [-1.44, 0.13] | .10 |
| Phase-out | -0.14 | [-0.71, 0.42] | .56 | -0.07 | [-0.39, 0.24] | .62 | -0.22 | [-1.01, 0.58] | .53 |
| Test Year | 0.49 | [-0.37, 1.33] | .24 | 0.41 | [-0.33, 1.18] | .28 | 0.52 | [-0.37, 1.44] | .25 |
| Standardized | | | | | | | | | |
| Phase-in | -0.04 | [-0.09, 0.01] | .07 | -0.02 | [-0.04, 0.00] | .10 | -0.08 | [-0.17, 0.01] | .08 |
| Phase-out | -0.01 | [-0.07, 0.05] | .64 | -0.00 | [-0.04, 0.03] | .76 | -0.02 | [-0.11, 0.07] | .61 |
| Test Year | 0.05 | [-0.06, 0.15] | .33 | 0.04 | [-0.05, 0.13] | .35 | 0.05 | [-0.06, 0.17] | .26 |
| Full Sample | | | | | | | | | |
| Original Scale | | | | | | | | | |
| Phase-in | -0.31 | [-0.55, -0.08] | .01 | -0.14 | [-0.24,-0.05] | .004 | -0.50 | [-0.99, -0.02] | .05 |
| Phase-out | -0.25 | [-0.68, 0.16] | .20 | -0.15 | [-0.42, 0.12] | .26 | -0.36 | [-1.02, 0.31] | .24 |
| Test Year | 0.32 | [-0.12, 0.78] | .14 | 0.29 | [-0.14, 0.71] | .15 | 0.33 | [-0.15, 0.83] | .15 |
| Standardized | | | | | | | | | |
| Phase-in | -0.03 | [-0.06, -0.01] | .009 | -0.01 | [-0.02,-0.00] | .005 | -0.06 | [-0.11, -0.01] | .03 |
| Phase-out | -0.02 | [-0.07, 0.02] | .26 | -0.01 | [-0.04, 0.01] | .33 | -0.04 | [-0.11, 0.04] | .27 |
| Test Year | 0.03 | [-0.02, 0.08] | .18 | 0.02 | [-0.02, 0.07] | .27 | 0.03 | [-0.02, 0.08] | .17 |

*Note.* Analysis on selective sample (n=203,142) included fixed effects of school by test type (n=7388) and year (n=13); 342 fixed effects singletons omitted; CIs and *p*s from wild cluster bootstrapping (999 replications; null imposed; School [n=2483] and year [n=13] as cluster variables. Analysis on full Ssample (n=2,152,909) included fixed effects of school by test type (n=7981) and year (n=13); 76 fixed effects singletons omitted; CIs and *p*s from wild cluster bootstrapping (999 replications; null imposed; school [n=2,667] and year [n=13] as cluster variables).

*Table S2. Demographic-adjusted regression coefficients (B), confidence intervals (CI) and p-values (p) for analyses of raw scores and standardized scores, Selective and Full Samples, 5th grade tests.*

| | Diminishing Impact | | | Number of fruit-years | | | Any fruit | | |
|---|---|---|---|---|---|---|---|---|---|
| | *B* | [95% CI] | *P* | *B* | [95% CI] | *p* | *B* | [95% CI] | *p* |
| Selective Sample | | | | | | | | | |
| Original Scale | | | | | | | | | |
| Phase-in | -0.39 | [-0.82, 0.01] | .06 | -0.18 | [-0.37, -0.02] | .07 | -0.68 | [-1.44, 0.10] | .07 |
| Phase-out | -0.16 | [-0.69, 0.39] | .55 | -0.08 | [-0.39, 0.23] | .57 | -0.25 | [-1.01, 0.59] | .48 |
| Test Year | 0.50 | [-0.45, 1.41] | .22 | 0.43 | [-0.31, 1.15] | .22 | 0.53 | [-0.37, 1.44] | .20 |
| Standardized | | | | | | | | | |
| Phase-in | -0.04 | [-0.09, 0.01] | .09 | -0.02 | [-0.04, 0.00] | .08 | -0.08 | [-0.17, 0.01] | .07 |
| Phase-out | -0.01 | [-0.07, 0.04] | .66 | -0.01 | [-0.04, 0.03] | .74 | -0.03 | [-0.13, 0.08] | .56 |
| Test Year | 0.05 | [-0.06, 0.16] | .29 | 0.04 | [-0.05, 0.13] | .34 | 0.06 | [-0.05, 0.16] | .25 |
| Full Sample | | | | | | | | | |
| Original Scale | | | | | | | | | |
| Phase-in | -0.29 | [-0.51, -0.07] | .02 | -0.13 | [-0.22,-0.05] | .005 | -0.48 | [-0.92, -0.04] | .04 |
| Phase-out | -0.23 | [-0.64, 0.18] | .23 | -0.13 | [-0.41, 0.12] | .28 | -0.33 | [-0.95, 0.29] | .26 |
| Test Year | 0.31 | [-0.11, 0.74] | .13 | 0.28 | [-0.14, 0.69] | .15 | 0.32 | [-0.13, 0.79] | .13 |
| Standardized | | | | | | | | | |
| Phase-in | -0.03 | [-0.05, -0.00] | .02 | -0.01 | [-0.02,-0.00] | .01 | -0.05 | [-0.10, -0.00] | .03 |
| Phase-out | -0.02 | [-0.06, 0.02] | .30 | -0.01 | [-0.04, 0.02] | .37 | -0.03 | [-0.10, 0.04] | .28 |
| Test Year | 0.03 | [-0.02, 0.07] | .18 | 0.02 | [-0.02, 0.07] | .27 | 0.03 | [-0.02, 0.08] | .17 |

*Note.* Controlling for sex, mother's and father's birth country (Norwegian, other Western, Non-Western), year by centrality, mother's and father's educations (7 levels), mother's and father's incomes and squared incomes. Analysis on selective sample (n=198,965) included fixed effects of school by test type (n=7376) and year (n=13); 349 fixed effects singletons omitted; CIs and *p*s from wild cluster bootstrapping (999 replications; null imposed; School [n=2480] and year [n=13] as cluster variables. Analysis on full Ssample (n=1,988,582) included fixed effects of school by test type (n=7969) and year (n=13); 76 fixed effects singletons omitted; CIs and *p*s from wild cluster bootstrapping (999 replications; null imposed; school [n=2,662] and year [n=13] as cluster variables).

*Table S3. Covariate-adjusted regression coefficients (B), confidence intervals (CI) and p-values (p) for analyses of the Selective and Full Samples, 5th grade test.*

| | Diminishing impact | | | Number of fruit-years | | | Any fruit | | |
|---|---|---|---|---|---|---|---|---|---|
| | *B* | [95% CI] | *p* | *B* | [95% CI] | *p* | *B* | [95% CI] | *p* |
| Selective Sample | | | | | | | | | |
| Original Scale | | | | | | | | | |
| Phase-in | -0.31 | [-0.78, 0.14] | .16 | -0.14 | [-0.34, 0.07] | .15 | -0.54 | [-1.30, 0.26] | .16 |
| Phase-out | -0.05 | [-0.71, 0.61] | .86 | -0.02 | [-0.41, 0.36] | .92 | -0.08 | [-1.14, 0.95] | .86 |
| Test Year | 0.40 | [-0.49, 1.27] | .34 | 0.33 | [-0.42, 1.12] | .36 | 0.42 | [-0.50, 1.38] | .34 |
| Standardized | | | | | | | | | |
| Phase-in | -0.04 | [-0.09, 0.02] | .18 | -0.01 | [-0.04, 0.01] | .17 | -0.07 | [-0.17, 0.04] | .17 |
| Phase-out | -0.00 | [-0.09, 0.08] | .93 | 0.00 | [-0.04, 0.05] | .96 | -0.01 | [-0.12, 0.11] | .86 |
| Test Year | 0.04 | [-0.07, 0.15] | .39 | 0.03 | [-0.06, 0.12] | .43 | 0.04 | [-0.06, 0.15] | .38 |
| Full Sample | | | | | | | | | |
| Original Scale | | | | | | | | | |
| Phase-in | -0.21 | [-0.38, -0.03] | .03 | -0.10 | [-0.16, -0.03] | .01 | -0.35 | [-0.75, 0.03] | .07 |
| Phase-out | -0.15 | [-0.53, 0.23] | .37 | -0.08 | [-0.31, 0.15] | .42 | -0.25 | [-0.77, 0.29] | .31 |
| Test Year | 0.16 | [-0.19, 0.54] | .33 | 0.15 | [-0.22, 0.52] | .41 | 0.17 | [-0.25, 0.58] | .35 |
| Standardized | | | | | | | | | |
| Phase-in | -0.02 | [-0.05, -0.00] | .05 | -0.01 | [-0.02,-0.00] | .04 | -0.04 | [-0.08, 0.00] | .05 |
| Phase-out | -0.01 | [-0.06, 0.03] | .44 | -0.01 | [-0.03, 0.02] | .54 | -0.03 | [-0.09, 0.04] | .36 |
| Test Year | 0.02 | [-0.02, 0.06] | .37 | 0.01 | [-0.03, 0.06] | .46 | 0.02 | [-0.02, 0.06] | .36 |

*Note.* Controlling for sex, mother's and father's birth country (Norwegian, other Western, Non-Western), year by centrality, mother's and father's educations (7 levels), mother's and father's incomes and squared incomes, size of cohort at school, size of cohort at school by treatment status, proportion of pupils exempted from the test, proportion exempted by treatment status (sex and birth countries not included in analysis on selective sample of non-immigrant boys). Analysis on Selective Sample (n=198,929) included fixed effects of school by test type (n=7373) and year (n=13); 349 fixed effects singletons were omitted; School (n=2479) and year (n=13) were cluster variables. Analysis on full sample (n=1,987,939) included fixed effects of school by test type (n=7966) and Year (n=13); 76 fixed effects singletons omitted; School (n=2,661) and year (n=13) were cluster variables. CIs and *p*s from wild cluster bootstrapping.

*Table S4. Example of covariate-adjusted analysis, full sample, 5th grade, standardized scores.*

| | *B* | [95 % CI] | p |
|---|---|---|---|
| Sex | | | |
| Male | Ref. | | |
| Female | -0.02 | [-0.051, 0.005] | .10 |
| Mother's Birth Country[1] | | | |
| Norway | Ref. | | |
| Western | 0.031 | [0.018, 0.045] | <.001 |
| Non-Western | -0.098 | [-0.119, 0.077] | .30 |
| Father's Birth Country[1] | | | |
| Norway | Ref. | | |
| Western | 0.031 | [0.018, 0.044] | <.001 |
| Non-Western | -0.098 | [-0.119, -0.077] | <.001 |
| Mother's Education | | | |
| Elementary | Ref. | | |
| Lower Secondary | 0.127 | [0.086, 0.169] | <.001 |
| Upper Secondary (basic) | 0.201 | [0.153, 0.250] | <.001 |
| Upper Secondary (completed) | 0.273 | [0.220,0.325] | <.001 |
| Higher Education (lower lever) | 0.458 | [0.403, 0.514] | <.001 |
| Higher Education (higher level) | 0.579 | [0.525, 0.632] | <.001 |
| PhD | 0.649 | [0.601, 0.696] | <.001 |
| Father's Education | | | |
| First Elementary | Ref. | | |
| Lower Secondary | 0.084 | [0.035, 0.133] | 0.003 |
| Upper Secondary (basic) | 0.159 | [0.110, 0.209] | <.001 |
| Upper Secondary (completed) | 0.205 | [0.153,0.256] | <.001 |
| Higher Education (lower lever) | 0.389 | [0.340, 0.439] | <.001 |
| Higher Education (higher level) | 0.486 | [0.435, 0.537] | <.001 |
| PhD | 0.556 | [0.498, 0.613] | <.001 |
| Mother's Income[2] | | | |
| Linear term | 0.028 | [0.024, 0.032] | <.001 |
| Quadratic term | -0.000 | [-0.001, -0.000] | <.001 |
| Father's Income[2] | | | |
| Linear term | 0.035 | [0.032, 0.038] | <.001 |
| Quadratic term | -0.001 | [-0.001, -0.001] | <.001 |
| Number of Fruit-Years[3] | | | |
| Phase-in | -0.013 | [-0.023, -0.003] | .013 |
| Phase-out | -0.011 | [-0.039, 0.017] | .39 |
| Fruit in Test Year[3] | 0.024 | [-0.022, 0.071] | .25 |

*Note.* The model included fixed effect of year (n=13) and school by test type (7966). Standard errors were clustered at year (n=13) and school (2661). [1]Western = EU/EEA, USA, Canada, Australia, and New Zealand . [2]Income is trimmed at 0 and standardized for each year. [3]Wild cluster bootstrapped confidence intervals reported for effect estimates (default two-way cluster-robust confidence intervals reported for other effects).

*Table S5. Descriptive statistics according to treatment group, 8th grade tests.*

| | Control | Treatment | Undetermined[a] |
|---|---|---|---|
| | *N* | *N* | *N* |
| Years | 12 | 12 | 12 |
| Schools | 1,841 | 1,109 | NA |
| School-Years | 20,936 | 8,685 | NA |
| Pupils | 496,058 | 139,038 | 104,177 |
| Observations | 1,470,989 | 410,157 | 301,163 |
| Missing test score | 53,094 | 14,197 | 12,240 |
| Non-standard tests | 42 | 45 | 0 |
| Valid standard tests | 1,417,855 | 385,923 | 288,923 |
| | *%* | *%* | *%* |
| Female | 49.2 | 49.7 | 49.3 |
| Non-immigrant | 77.0 | 76.7 | 80.2 |
| Higher education, mother | 48.5 | 47.2 | 40,6 |
| Higher education, father | 37.2 | 31.2 | 31.4 |
| | *Mean (SD)* | *Mean (SD)* | *Mean (SD)* |
| Mother's income[b] | 370' (273') | 343' (247') | 304' (220') |
| Father's income[b] | 579' (539') | 515' (429') | 484' (403') |
| Centrality[c] | 836 (109) | 757 (162) | 873 (102) |
| Test scores | | | |
| Mathematics | 28.6 (11.5) | 27.9 (11.4) | 29.4 (11.9) |
| English | 28.5 (11.8) | 28.1 (11.9) | 24.6 (10.2) |
| Reading | 26.4 (8.9) | 26.0 (8.9) | 27.3 (9.0) |
| Standardized scores | | | |
| Mathematics | 0.02 (1.00) | -0.04 (0.99) | -0.03 (1.00) |
| English | 0.01 (1.00) | -0.02 (1.01) | -0.02 (1.01) |
| Reading | 0.01 (1.00) | -0.03 (1.00) | -0.02 (1.00) |

*Note.* The table excludes data from 2007 where treatment status for all data is Undetermined (60,037 pupils and 172,853 observations). [a]The parameters for the Undetermined category are dominated by data from the earliest years (70% of the observations are from 2008-2009). [b]Income reported in NOK 1000; negative values trimmed at 0. [c]Urbanicity of the schools' municipalities, maximum = 1000. Abbreviations: NA= Not available; SD = Standard Deviation.

*Table S6. Regression coefficients (B), confidence intervals (CI) and p-values (p) for analyses of 8th grade tests (no data in 2007).*

| | Diminishing impact | | | Number of fruit-years | | |
|---|---|---|---|---|---|---|
| | *B* | [95% CI] | *P* | *B* | [95% CI] | *P* |
| Selective Sample | | | | | | |
| Original Scale | | | | | | |
| Phase-in | -1.04 | [-1.88, -0.19] | .02 | -0.17 | [-0.32, -0.03] | .02 |
| Phase-out | -1.01 | [-1.90, -0.16] | .03 | -0.16 | [-0.35, 0.04] | .10 |
| Standardized | | | | | | |
| Phase-in | -0.10 | [-0.17, -0.02] | .01 | -0.02 | [-0.02, -0.00] | .01 |
| Phase-out | -0.10 | [-0.19, -0.00] | .04 | -0.01 | [-0.03, 0.00] | .10 |
| Full Sample | | | | | | |
| Original Scale | | | | | | |
| Phase-in | -0.39 | -1.01, 0.21 | .18 | -0.07 | -0.14,0.01 | .08 |
| Phase-out | -0.47 | -1.18, 0.23 | .15 | -0.10 | -0.23, 0.03 | .10 |
| Standardized | | | | | | |
| Phase-in | -0.03 | -0.08, 0.02 | .21 | -0.01 | -0.01, 0.00 | .16 |
| Phase-out | -0.04 | -0.10, 0.02 | .16 | -0.01 | -0.02, 0.00 | .10 |

*Note.* Selective Sample: Fixed effects of school by test type (n=7214) and year (n=12), clustered at school (n=2,423) and year (n=12), observations = 174,510. 517 fixed effects singletons omitted. Full sample: Fixed effects of school by test type (n=8334) and year (n=12), clustered at school (n=2801) and year (n=12), observations = 1,803,394. Bootstrapped, with null imposed, 999 replications. 384 fixed effects singletons omitted. There were 79,878 missing observations (exempted/not participated) 45,764 in control (3.1%) and 22,043 in treatment (5.4%). Apart from data on treatment status missing entirely for the year 2007, there were 289,092 observations with missing on treatment status (ranging from 2057 in 2017 to 144921 in 2009). The effect of having fruit in the test year and the impact model of having any fruit cannot be properly estimated for the 8th grade test (both treatment and control received fruit in the test year during the policy and all pupils in the treatment schools had received fruit at least one year).

*Table S7. Regression coefficients (B), confidence intervals (CI) and p-values (p) for alternative analysis of 8th grade data (using current school as a proxy for elementary school treatment status).*

| | Diminishing Impact | | | Number of fruit-years | | | Any fruit | | |
|---|---|---|---|---|---|---|---|---|---|
| | *B* | [95% CI] | *P* | *B* | [95% CI] | *p* | *B* | [95% CI] | *p* |
| Selective Sample | | | | | | | | | |
| Original Scale | | | | | | | | | |
| Phase-in | -0.01 | [-0.64, 0.63] | .98 | -0.05 | [-0.21, 0.10] | .43 | 0.36 | [-0.35, 1.07] | .32 |
| Phase-out | -0.00 | [-0.65, 0.65] | .99 | -0.03 | [-0.20, 0.13] | .62 | 0.32 | [-0.28, 0.92] | .25 |
| Standardized | | | | | | | | | |
| Phase-in | -0.00 | [-0.07, 0.06] | .89 | -0.01 | [-0.02, 0.01] | .38 | -0.03 | [-0.04, 0.10] | .35 |
| Phase-out | -0.00 | [-0.06, 0.06] | .95 | -0.00 | [-0.02, 0.01] | .61 | -0.03 | [-0.03, 0.08] | .30 |
| Full Sample | | | | | | | | | |
| Original Scale | | | | | | | | | |
| Phase-in | 0.02 | [-0.28, -0.32] | .86 | -0.03 | [-0.12, 0.06] | .47 | 0.15 | [-0.32, 0.63] | .53 |
| Phase-out | -0.04 | [-0.28, 0.20] | .73 | -0.04 | [-0.13, 0.05] | .29 | 0.03 | [-0.34, 0.41] | .86 |
| Standardized | | | | | | | | | |
| Phase-in | -0.00 | [-0.03, 0.02] | .99 | -0.00 | [-0.01,-0.00] | .42 | 0.01 | [-0.03, 0.05] | .76 |
| Phase-out | -0.01 | [-0.03, 0.01] | .43 | -0.01 | [-0.01, 0.00] | .22 | -0.01 | [-0.04, 0.02] | .66 |

Note. Selective Sample: Fixed effects of school by test type (n=3669) and year (n=13), clustered at school (n=1,235) and year (n=13), observations = 229,011. Bootstrapped, with null imposed, 999 replications. 140 fixed effects singletons omitted. Full sample: Fixed effects of school by test type (n=3927) and year (n=13), clustered at school (n=1,318) and year (n=13), observations = 2,224,169. Bootstrapped, with null imposed, 999 replications. 40 fixed effects singletons omitted.

*Table S8. Regression coefficients (B), confidence intervals (CI) and p-values (p) for analyses of 10th grade exams when Phase-out includes years 2015-2019 (exposure in the past but not the previous year after to policy after policy was repealed).*

| | Diminishing impact | | | Number of fruit-years | | | Any fruit | | |
|---|---|---|---|---|---|---|---|---|---|
| | *B* | [95% CI] | *P* | *B* | [95% CI] | *P* | *B* | [95% CI] | *p* |
| Phase-In | -0.19 | [-0.41, 0.03] | 0.08 | -0.13 | [-0.24, -0.02] | .03 | -0.23 | [-0.59, 0.13] | 0.19 |
| Phase-Out | -0.27 | [-0.40, -0.10] | 0.007 | -0.10 | [-0.15, -0.04] | .002 | -0.50 | [-0.83, -0.18] | 0.008 |

*Note.* Scaled as "school points" (grade multiplied by 10). Fixed effects of school by test type (n=2707) and year (n=18), clustered at school (n=954) and year (n=18), observations = 27,050. CIs and ps based on Wild Cluster Bootstrapping, null imposed, 999 replications. 216 fixed effects singletons omitted. Phase-In from Phase-Out from 2015 to 2019

*Table S9. Regression coefficients (B), confidence intervals (CI) and p-values (p) for analyses of 10th grade exams when Phase-out is coded as decreasing years of exposure (years 2018-2019)*

| | Diminishing impact | | | Number of fruit-years | | | Any fruit | | |
|---|---|---|---|---|---|---|---|---|---|
| | *B* | [95% CI] | *P* | *B* | [95% CI] | *P* | *B* | [95% CI] | *p* |
| Phase-In | -0.28 | [-0.43, -0.12] | 0.003 | -0.12 | [-0.17, -0.06] | .001 | -0.40 | [-0.71, -0.09] | 0.02 |
| Phase-Out | -0.17 | [-0.40, 0.07] | 0.12 | -0.06 | [-0.14, 0.02] | .11 | -0.29 | [-0.71, 0.15] | 0.18 |

*Note.* Fixed effects of school by test type (n=2707) and year (n=18), clustered at school (n=954) and year (n=18), observations = 27,050. CIs and ps based on Wild Cluster Bootstrapping, null imposed, 999 replications. 216 fixed effects singletons omitted.

*Table S10. Regression coefficients (B), confidence intervals (CI) and p-values (p) for analyses of 9th grade national tests.*

| | Diminishing impact | | | Number of fruit-years | | |
|---|---|---|---|---|---|---|
| | *B* | [95% CI] | *P* | *B* | [95% CI] | *P* |
| Selective Sample | | | | | | |
| Original Scale | | | | | | |
| Phase-in | -1.46 | [-3.11, 0.20] | .08 | -0.18 | [-0.38, 0.02] | .09 |
| Phase-out | -1.53 | [-3.32, 0.23] | .08 | -0.19 | [-0.37, -0.00] | .04 |
| Standardized | | | | | | |
| Phase-in | -0.14 | [-0.30, 0.02] | .09 | -0.02 | [-0.04, 0.00] | .08 |
| Phase-out | -0.15 | [-0.31, 0.02] | .08 | -0.02 | [-0.04, 0.00] | .06 |
| Full Sample | | | | | | |
| Original Scale | | | | | | |
| Phase-in | -1.19 | -1.86, -0.53 | .002 | -0.10 | -0.18, -0.02 | .02 |
| Phase-out | -1.29 | -2.07, -0.50 | .001 | -0.13 | -0.21, -0.06 | .006 |
| Standardized | | | | | | |
| Phase-in | -0.11 | -0.17, -0.05 | .002 | -0.01 | -0.02, 0.00 | .05 |
| Phase-out | -0.12 | -0.18, -0.06 | <.001 | -0.01 | -0.02, -0.00 | .007 |

*Note.* Selective Sample: Fixed effects of school by test type (n=2319) and year (n=10), clustered at school (n=1,167) and year (n=10), observations = 111,728. Bootstrapped, with null imposed, 999 replications. 99 fixed effects singletons omitted. Full sample: Fixed effects of school by test type (n=2495) and year (n=10), clustered at school (n=1252) and year (n=10), observations = 1,140,858. Bootstrapped, with null imposed, 999 replications. 19 fixed effects singletons omitted.

*Table S11. Associations between differences in fruit exposure and aggregated differences in test scores (yearly mean differences between control and treatment), regardless of phase-in/phase-out.*

| | Diminishing impact | | | Number of fruit-years | | | Any fruit | | |
|---|---|---|---|---|---|---|---|---|---|
| | *B* | [95% CI] | *P* | *B* | [95% CI] | *P* | *B* | [95% CI] | *p* |
| Original Scale | | | | | | | | | |
| 5th grade | -0.31 | [-0.50, -0.11] | .005 | -0.14 | [-0.24, -0.05] | .006 | -0.49 | [-0.86, -0.13] | 0.01 |
| 8th grade | -0.38 | [-0.82, 0.06] | .08 | -0.07 | [-0.14, -0.01] | .02 | NA | NA | NA |
| 8th grade (alt.) | 0.05 | [-0.18, 0.29] | .62 | -0.02 | [-0.08, 0.04] | .53 | 0.20 | [-0.26, 0.66] | 0.36 |
| 9th grade | -1.35 | [-2.14, -0.55] | .005 | -0.12 | [-0.21, -0.03] | .01 | NA | NA | NA |
| 10th grade | -0.27 | [-0.41, -0.13] | .001 | -0.11 | [-0.15, -0.06] | <.001 | -0.44 | [-0.73, -0.15] | .005 |
| All | -0.30 | [-0.40, -0.19] | <.001 | -0.11 | [-0.14, -0.08] | <.001 | -0.46 | [-0.68, -0.24] | <.001 |
| All (w/8th alt.) | -0.24 | -0.37, -0.11 | <.001 | -0.09 | [-0.13, -0.06] | <.001 | -0.38 | -0.63, -0.13 | .005 |
| Standardized | | | | | | | | | |
| 5th grade | -0.03 | [-0.05, -0.01] | .003 | -0.02 | [-0.03, -0.01] | .005 | -0.06 | [-0.09, -0.02] | 0.01 |
| 8th grade | -0.03 | [-0.07, 0.01] | .11 | -0.01 | [-0.01, 0.00] | .06 | NA | [NA] | NA |
| 8th grade (alt.) | 0.00 | [-0.02, 0.02] | .90 | -0.00 | [-0.01, 0.00] | .48 | 0.01 | [-0.03, 0.05] | 0.65 |
| 9th grade | -0.13 | [-0.21, -0.04] | .01 | -0.01 | [-0.02, -0.00] | .02 | NA | [NA] | NA |
| 10th grade | -0.03 | [-0.04, -0.01] | .001 | -0.01 | [-0.02, -0.01] | <.001 | -0.04 | [-0.07, -0.02] | .005 |
| All | -0.03 | [-0.04, -0.02] | <.001 | -0.01 | [-0.01, -0.01] | <.001 | -0.05 | [-0.07, -0.03] | <.001 |
| All (w/8th alt.) | -0.03 | [-0.04, -0.01] | <.001 | -0.01 | [-0.01, -0.01] | <.001 | -0.04 | [-0.07, -0.02] | .002 |

Note "All" includes grade indicators and clustering on cohort (n=106, cluster n=23). No individual-level standard deviations were available for 10th grade, therefore the standardized scores were approximated by using grades (school points divided by 10) instead of school points (this approximates a standardized scaling because the standard deviations in national test results with similar means were close to 10).

*Table S12. Within-subject analyses for cohorts 2001 to 2004.*

| | Diminishing impact | | | Number of fruit-years | | |
|---|---|---|---|---|---|---|
| | *B* | [95% CI] | *p* | *B* | [95% CI] | *p* |
| Selective Sample | | | | | | |
| Original Scale | -2.45 | [-6.74, 1.84] | .26 | -0.11 | [-0.27, 0.05] | .18 |
| Standardized | -0.12 | [-0.57, 0.34] | .62 | -0.003 | [-0.02, 0.01] | .70 |
| Full Sample | | | | | | |
| Original Scale | -2.45 | [-4.65, -0.24] | .03 | -0.12 | [-0.20, -0.03] | .007 |
| Standardized | -0.23 | [-0.48, 0.02] | .08 | -0.008 | [-0.017, 0.002] | .11 |

*Note.* Selective Sample: Observations 152,768, Fixed effects (56,890) of person (21,397) by test-type (3); 7,471 singleton observations omitted. Full sample: Observations 1,635,564, Fixed effects (607,736) of person (224,338) by test-type (3); 69,707 singleton observations omitted. Cluster-robust standard errors clustered at School in 5$^{th}$ grade (N= 2,404 full sample and N=2,237 for selective sample). The effect estimates represent the interaction of the change from 5$^{th}$ grade scores to 8$^{th}$ and 9$^{th}$ with the level of exposure to free fruit in elementary school (controlled for the changes of each cohort and the overall difference of change for the treatment group).

*Table S13. Within-subject analyses for cohorts 1997 to 2006*

| | Diminishing impact | | | Number of fruit-years | | |
|---|---|---|---|---|---|---|
| | *B* | [95% CI] | *p* | *B* | [95% CI] | *p* |
| Selective Sample | | | | | | |
| Original Scale | -0.72 | [-1.65, 0.21] | .11 | -0.07 | [-0.16, 0.02] | .10 |
| Standardized | -0.03 | [-0.14, 0.10] | .60 | -0.002 | [-0.012, 0.009] | .72 |
| Full Sample | | | | | | |
| Original Scale | -0.52 | [-1.27, 0.20] | .13 | -0.06 | [-0.12, 0.01] | .08 |
| Standardized | -0.003 | [-0.05, 0.04] | .86 | -0.001 | [-0.006, 0.004] | .66 |

*Note.* Selective Sample: Observations 423,085, Fixed effects (163,397) of person (58,425) by test-type (3); 13,053 singleton observations omitted. Full sample: Observations 4,179,661, Fixed effects (1,616,951) of person (570,769) by test-type (3); 110,223 singleton observations omitted. Cluster-robust standard errors clustered at School in 5th grade (n= 2,651 full sample and n=2,542 for selective sample) and cohort (n=10). The effect estimates represent the interaction of the change from 5th grade scores to 8th and 9th with the level of exposure to free fruit in elementary school (controlled for the changes of each cohort and the overall difference of change for the treatment group).

*Table S14. Regression coefficients (B), confidence intervals (CI) and p-values (p) for analyses of Selective and Full Samples, 5th grade tests, excluding the year 2019 (original pre-registered analyses).*

| | Diminishing impact | | | Number of fruit-years | | | Any fruit | | |
|---|---|---|---|---|---|---|---|---|---|
| | *B* | [95% CI] | *p* | *B* | [95% CI] | *p* | *B* | [95% CI] | *p* |
| Selective Sample | | | | | | | | | |
| Raw score | | | | | | | | | |
| Phase-in | -0.34 | [-0.83, 0.14] | .15 | -0.16 | [-0.35, 0.03] | .10 | -0.57 | [-1.54, 0.43] | .20 |
| Phase-out | -0.03 | [-0.70, 0.65] | .91 | -0.01 | [-0.39, 0.36] | .93 | -0.03 | [-0.99, 0.91] | .94 |
| Test Year | 0.59 | [-0.51, 1.74] | .27 | 0.52 | [-0.38, 1.49] | .24 | 0.62 | [-0.58, 1.86] | .27 |
| Standardized | | | | | | | | | |
| Phase-in | -0.04 | [-0.09, 0.02] | .16 | -0.02 | [-0.04, 0.01] | .17 | -0.07 | [-0.19, 0.06] | .22 |
| Phase-out | 0.00 | [-0.08, 0.08] | .98 | 0.00 | [-0.04, 0.04] | .87 | -0.00 | [-0.11, 0.11] | .98 |
| Test Year | 0.06 | [-0.07, 0.18] | .29 | 0.05 | [-0.05, 0.15] | .28 | 0.06 | [-0.08, 0.21] | .26 |
| Full Sample | | | | | | | | | |
| Raw score | | | | | | | | | |
| Phase-in | -0.24 | [-0.44, -0.04] | .02 | -0.12 | [-0.20, -0.04] | .01 | -0.36 | [-0.78, 0.04] | .07 |
| Phase-out | -0.09 | [-0.34, 0.14] | .37 | -0.06 | [-0.20, 0.08] | .34 | -0.10 | [-0.40, 0.19] | .41 |

| | | | | | | | | | |
|---|---|---|---|---|---|---|---|---|---|
| Test Year | 0.45 | [-0.05, 0.93] | .07 | 0.42 | [0.00, 0.82] | .05 | 0.46 | [-0.07, 0.97] | .07 |
| Standardized | | | | | | | | | |
| Phase-in | -0.02 | [-0.05, -0.00] | .03 | -0.01 | [-0.02,-0.00] | .02 | -0.04 | [-0.09, 0.00] | .05 |
| Phase-out | -0.01 | [-0.02, 0.01] | .57 | -0.00 | [-0.01, 0.01] | .61 | -0.01 | [-0.04, 0.02] | .53 |
| Test Year | 0.04 | [-0.00, 0.09] | .06 | 0.04 | [-0.00, 0.08] | .06 | 0.05 | [-0.00, 0.10] | .07 |

*Note.* Analysis on Selective Sample (n=191,720) included fixed effects of school by test type (n=7359) and year (n=12); 353 fixed effects singletons omitted; CIs and *p*s from wild cluster bootstrapping (999 replications; null imposed; School [n=2472] and year [n=12] as cluster variables. Analysis on full sample (n=1,986,945) included fixed effects of school by test type (n=7969) and year (n=12); 71 fixed effects singletons omitted; CIs and *p*s from wild cluster bootstrapping; school [n=2663] and year [n=12] as cluster variables).

*Table S15. Results from Linear Mixed Models, $5^{th}$ grade tests.*

| | Diminishing impact | | | Number of fruit-years | | | Any fruit | | |
|---|---|---|---|---|---|---|---|---|---|
| | *B* | [95% CI] | *P* | *B* | [95% CI] | *P* | *B* | [95% CI] | *p* |
| Selective Sample | | | | | | | | | |
| Phase-in | -0.42 | [-0.67, -0.18] | 0.01 | -0.19 | [-0.30, -0.08] | .01 | -0.71 | [-1.22, -0.19] | 0.03 |
| Phase-out | -0.19 | [-0.54, 0.16] | 0.31 | -0.10 | [-0.32, 0.13] | .41 | -0.29 | [-0.87, 0.28] | 0.34 |
| Test Year | 0.54 | [0.10, 0.98] | 0.04 | 0.47 | [0.05, 0.89] | .06 | 0.55 | [0.03, 1.08] | 0.07 |
| Full Sample | | | | | | | | | |
| Phase-in | No convergence | | | -0.14 | [-0.22, -0.06] | .007 | -0.49 | [-0.89, -0.09] | .04 |
| Phase-out | No convergence | | | -0.17 | [-0.32, -0.02] | .06 | -0.40 | [-0.83, 0.02] | .10 |
| Test Year | No convergence | | | 0.32 | [0.02, 0.62] | .06 | 0.36 | [-0.04, 0.76] | .11 |

Abbreviations: CI = (Wald) Confidence Intervals. B = unstandardized regression coefficient.

*Table S16. Example of regression results from a Linear Mixed Model of all 5th grade tests, linear impact variable.*

| Fixed Effect | Coef | SE | Df | t | *p* |
|---|---|---|---|---|---|
| Constant | 24.33 | 0.95 | 9.37 | 82.35 | <.001 |
| Treatment | -1.06 | 0.15 | 17.73 | -7.19 | <.001 |
| Phase-in | 0.33 | 0.10 | 8.99 | 3.45 | 0.007 |
| Phase-out | 0.29 | 0.18 | 8.99 | 1.58 | 0.15 |
| Phase-in*Treatment | -0.14 | 0.04 | 9.12 | -3.48 | 0.007 |
| Phase-out*Treatment | -0.17 | 0.08 | 9.00 | -2.17 | 0.06 |
| Reading | -6.59 | 0.01 | 1,403,016.63 | -571.00 | <.001 |
| Mathematics | -0.02 | 0.01 | 1,405,621.97 | -1.59 | 0.11 |
| Reading: Treatment | 0.33 | 0.03 | 1,236,477.75 | 12.90 | <.001 |
| Mathematics*Treatment | 0.51 | 0.03 | 1,247,251.88 | -19.90 | <.001 |
| Test Year Fruit | 0.58 | 0.36 | 8.99 | 1.64 | 0.14 |
| Test Year Fruit*Treatment | 0.32 | 0.15 | 9.17 | 2.12 | 0.06 |
| Random Effects | N | | SD | | |
| Pupil (intercept) | 760,234 | | 5.62 | | |
| School (intercept) | 2,691 | | 1.69 | | |
| Time (intercept) | 13 | | 0.50 | | |
| Time (Treatment slope) | 13 | | 0.18 (intercept-slope corr.: -0.63) | | |
| School*Time (intercept) | 29,437 | | 1.69 | | |
| Residual | 2,153,978 | | 6.03 | | |

*Table S17. Coding of the Variables Used for the three different impact models.*

| | 2007 | 2008 | 2009 | 2010 | 2011 | 2012 | 2013 | 2014 | 2015 | 2016 | 2017 | 2018 | 2019 |
|---|---|---|---|---|---|---|---|---|---|---|---|---|---|
| 5th grade | | | | | | | | | | | | | |
| Fruit-Years (Phase-in) | 0 | 1 | 2 | 3 | 4 | 4 | 4 | 4 | 0 | 0 | 0 | 0 | 0 |
| Fruit-Years (Phase-out) | 0 | 0 | 0 | 0 | 0 | 0 | 0 | 0 | 3 | 2 | 1 | 0 | 0 |
| Fruit in Test Year | 1 | 1 | 1 | 1 | 1 | 1 | 1 | 0 | 0 | 0 | 0 | 0 | 0 |
| Non-linear (Phase-in) | 0 | 1 | 1.5 | 1.75 | 1.88 | 1.88 | 1.88 | 1.88 | 0 | 0 | 0 | 0 | 0 |
| Non-linear (Phase-out) | 0 | 0 | 0 | 0 | 0 | 0 | 0 | 0 | 1.75 | 1.5 | 1 | 0 | 0 |
| Non-linear (Phase-out Alt.) | 0 | 0 | 0 | 0 | 0 | 0 | 0 | 0 | 0.88 | 0.38 | 0.13 | 0 | 0 |
| Any Fruit (Phase-in) | 0 | 1 | 1 | 1 | 1 | 1 | 1 | 1 | 0 | 0 | 0 | 0 | 0 |
| Any fruit (Phase-out) | 0 | 0 | 0 | 0 | 0 | 0 | 0 | 0 | 1 | 1 | 1 | 0 | 0 |
| 8th grade | | | | | | | | | | | | | |
| Fruit-Years (Phase-in) | NA | 1 | 2 | 3 | 4 | 5 | 6 | 7 | 0 | 0 | 0 | 0 | 0 |
| Fruit-Years (Phase-out) | NA | 0 | 0 | 0 | 0 | 0 | 0 | 0 | 6 | 5 | 4 | 3 | 2 |
| Non-linear (Phase-in) | NA | 1 | 1.5 | 1.75 | 1.88 | 1.94 | 1.97 | 1.98 | 0 | 0 | 0 | 0 | 0 |
| Non-linear (Phase-out) | NA | 0 | 0 | 0 | 0 | 0 | 0 | 0 | 1.97 | 1.94 | 1.89 | 1.75 | 1.5 |

*Note.* The Fruit-Years variables indicate the number of years the pupils received free fruit in the school-years preceeding the test (i.e., the learning years). The Fruit in Test Year variable indicate whether the pupils received fruit in the year the test took place (the test is in the beginning of the school year). The Non-linear Impact variables reflect the authors' a priori best guess of the potential impact of number of years of free fruit (diminishing impact of additional years). Abbreviations: NA = Not available (data for past year's school missing);

*Table S18. Bayes Factors comparing null effects against a distribution of positive effect sizes, 5th grade.*

| | Diminishing impact | Number of fruit-years | Any fruit |
|---|---|---|---|
| Selective Sample | 0.14 | 0.12 | 0.21 |
| Full Sample | 0.06 | 0.03 | 0.11 |

*Table S19. Regression coefficients (B), confidence intervals (CI) and p-values (p) for analyses of the Selective and Full Samples when controlling for municipality-specific year effects, 5th grade tests.*

| | Non-linear impact | | | Number of fruit-years | | | Any fruit | | |
|---|---|---|---|---|---|---|---|---|---|
| | *B* | [95% CI] | *P* | *B* | [95% CI] | *p* | *B* | [95% CI] | *p* |
| Selective Sample | | | | | | | | | |
| Phase-in | -0.37 | [-0.60, -0.14] | .004 | -0.16 | [-0.28, -0.05] | .009 | -0.63 | [-1.02, -0.23] | .005 |
| Phase-out | -0.07 | [-0.38, 0.24] | .63 | -0.03 | [-0.21, 0.15] | .74 | -0.12 | [-0.60, 0.36] | .61 |
| Test year | 0.40 | [-0.03, 0.84] | .07 | 0.32 | [-0.09, 0.74] | .11 | 0.43 | [-0.01, 0.87] | .052 |
| Full Sample | | | | | | | | | |
| Phase-in | -0.32 | [-0.48, -0.16] | .001 | -0.14 | [-0.21, -0.07] | .001 | -0.54 | [-0.84, -0.24] | .002 |
| Phase-out | -0.22 | [-0.51, 0.08] | .14 | -0.11 | [-0.29, 0.06] | .19 | -0.35 | [-0.80, 0.09] | .11 |
| Test year | 0.35 | [0.07, 0.63] | .02 | 0.31 | [0.02, 0.61] | .04 | 0.36 | [0.07, 0.65] | .02 |

*Note.* Analysis on Selective Sample (n=203,112) included fixed effects of School by Test Type (n=7388) and Year by School Municipality (n=5242); 372 fixed effects singletons omitted; CIs and *p*s from default robust standard errors clustered on School (n=2483) and Year (n=13). Analysis on Full Sample (n=2,152,903) included fixed effects of School by Test Type (n=7981) and Year by School Municipality (n=5570); 69 fixed effects singletons omitted; CIs and *p*s from default robust standard errors clustered on School (n=2,667) and Year (n=13).

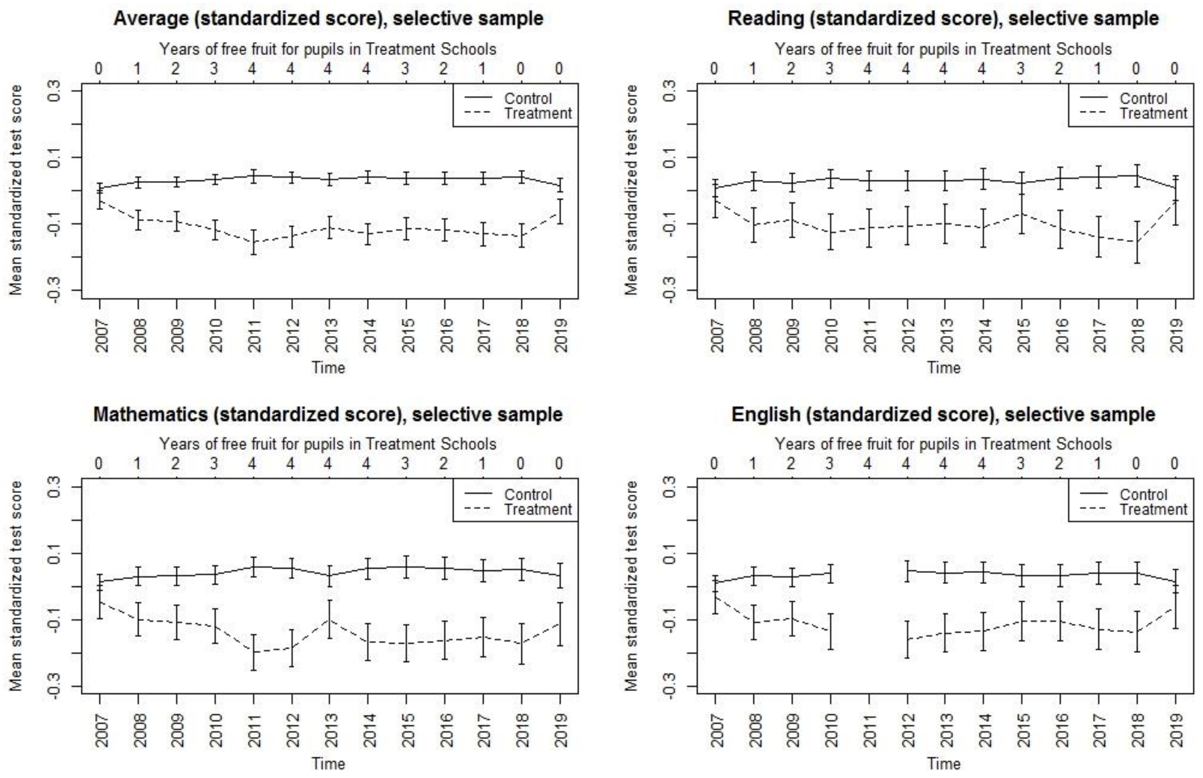


**Figure S1**. Standardized scores, selective sample, 5th grade tests. Error bars are 95% confidence intervals of means.

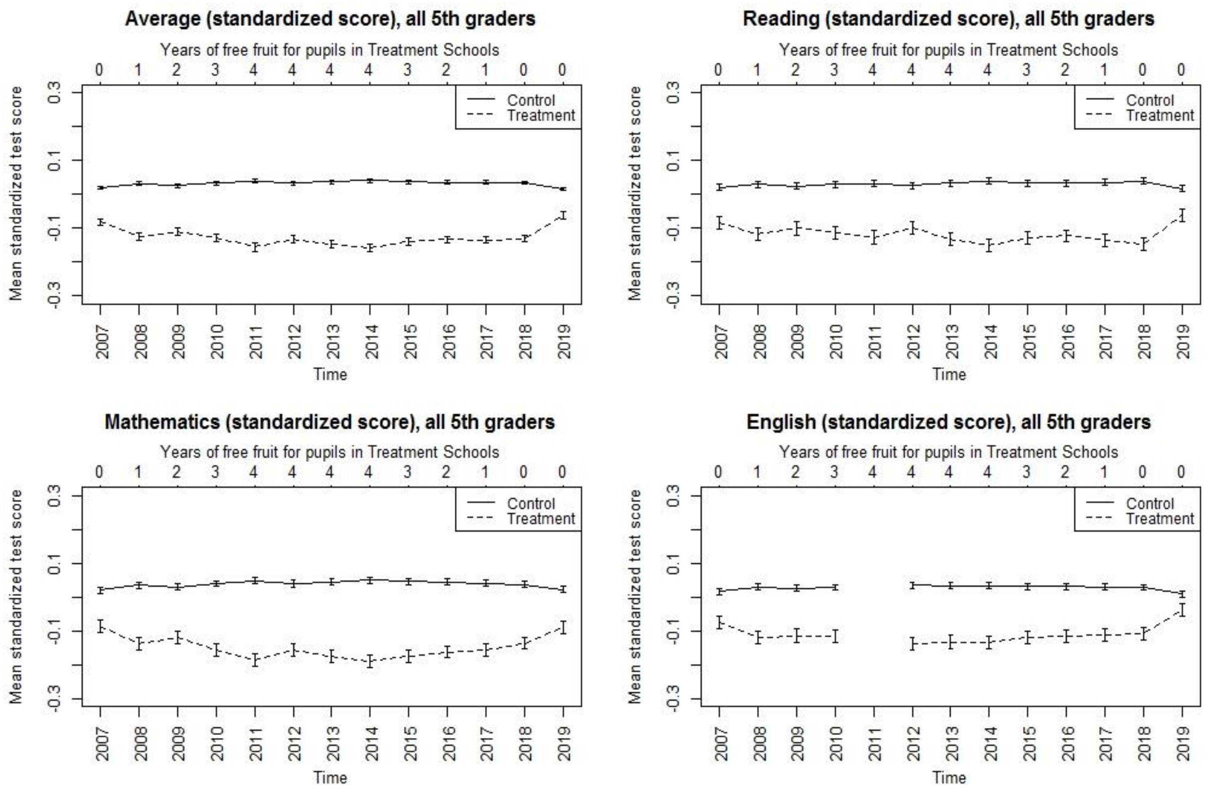


**Figure S2.** Standardized scores, full sample, 5th grade tests. Error bars are 95% confidence intervals of means.
